# Covid-19 pandemic lessons: Uncritical communication of test results can induce more harm than benefit and raises questions on standardized quality criteria for communication and liability

**DOI:** 10.1101/2020.11.20.20235424

**Authors:** Franz Porzsolt, Gerit Pfuhl, Robert M Kaplan, Martin Eisemann

## Abstract

**Background:** The COVID-19 pandemic is characterized by both health and economic risks. A ‘safety loop’ model postulates risk-related decisions are not based on objective and measurable risks but on the subjective perception of those risks. We here illustrate a quantification of the difference between objective and subjective risks.

**Method:** The objective risks (or chances) can be obtained from traditional 2 × 2 tables by calculating the positive (+LR) and negative (-LR) likelihood ratios. The subjective perception of objective risks is calculated from the same 2 × 2 tables by exchanging the X- and Y-axes. The traditional 2 × 2 table starts with the hypothesis, uses a test and a gold standard to confirm or exclude the investigated condition. The 2 × 2 table with inverted axes starts with the communication of a test result and presumes that the communication of bad news (whether right or false) will induce ‘perceived anxiety’ while good news will induce ‘perceived safety’. Two different functions (confirmation and exclusion) of both perceptions (perceived anxiety and safety) can be quantified with those calculations.

**Results:** The analysis of six published tests and of one incompletely reported test on COVID-19 polymerase chain reactions (completed by four assumptions on high and low sensitivities and specificities) demonstrated that none of these tests induces ‘perceived safety’. Eight of the ten tests confirmed the induction of perceived anxiety with +LRs (range 3.1 – 5900). In two of these eight tests a -LR (0.25 and 0.004) excluded the induction of perceived safety.

**Conclusions:** Communication of test results caused perceived anxiety but not perceived safety in 80% of the investigated tests. Medical tests – whether right or false – generate strong psychological messages. In the case of COVID-19 tests may induce more perceived anxiety than safety.

## Introduction

In February 2020 it became clear to epidemiologists that the new SARS-CoV-2 had travelled around the globe, and subsequently on 11^th^ of March 2020 the WHO classified COVID-19 as a pandemic (1).

Seven months later, the COVID-19 pandemic continues to advance in many countries. In the absence of reliable data and tools politicians had to make decisions based on limited information. Accordingly, there is considerable variation by country on strategies to control the number of infected persons. Some countries imposed complete lockdowns whereas other countries issued public health advice and partial lockdowns. Compliance with public health advice rests on a citizen’s subjective probability of contagion, the subjective assessment on the noxiousness of contagion, and the confidence in the real-world effectiveness of the recommended interventions (2). The higher the perceived efficacy of e.g. imposed COVID-19 restrictions the better was the mental health among participants in a recent observational online study in six countries (3).

As we learn on the go about SARS-CoV-2 High Reliability Organizations (HRO) should embrace the uncertainty to successfully cope with risk in sophisticated and complex systems (4). The four recommendations to High Reliability Organizations (HROs) require a) recognition of the difference between reliability and safety, b) training of members of HROs to provide appropriate responses to crisis situations, c) usage of sophisticated forms of organizational learning, and d) to use redundancy extensively.

In line with those recommendations a safety loop has been proposed (Fig. 1). The ‘safety loop’ describes the interrelationship between objective risks, risk communication, the resulting subjective perception of objective risks, the derived consequences from the subjective perception of the objective risk and finally the effect of the decisions onto the objective risk (5, 6).

**Fig. 1:**
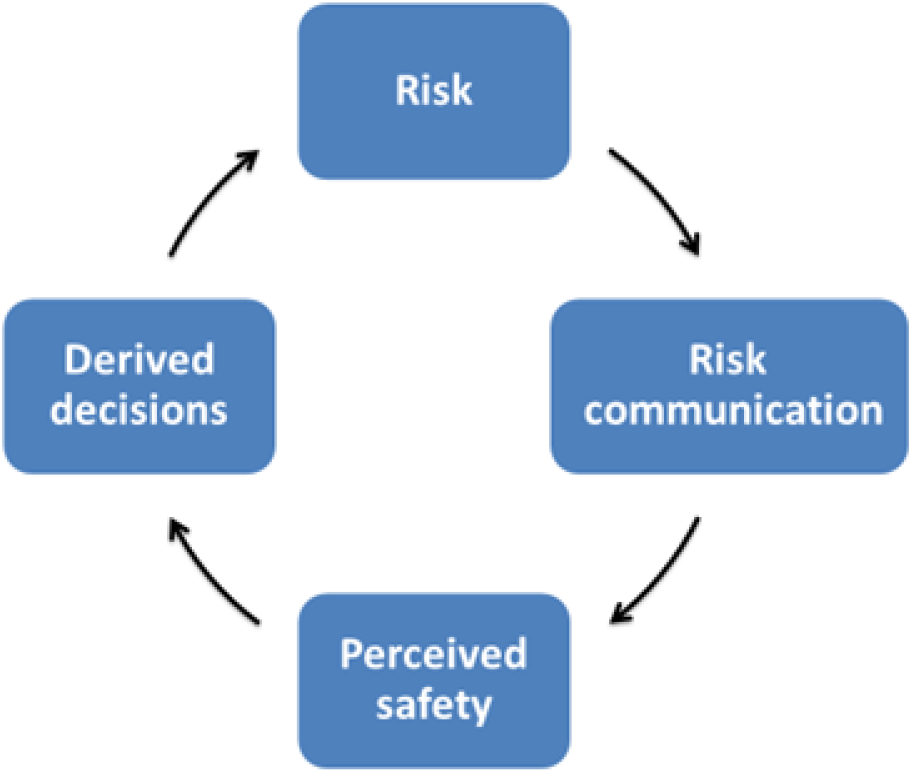
The Safety Loop. The safety loop describes the association and the mutual influence of an objective risk and the subjective perception of the objective risk (perceived safety). Objective risks can be assessed as the incidence of event times the size of damage (probability by noxiousness). The subjective perception of the objective risks can be described either by psychometric methods (suppl. I, 28 – 32) or may be expressed by odds ratios (perceived safety or perceived anxiety) as described in this paper. Explanation of the safety loop: Existing risks trigger the risk communication. The risk communication affects the subjective perception of objective risks. The subjective perception of the risk (perceived safety or anxiety) depends not only on communication but several factors (7) that will govern the derived decision. The loop shows that a high-risk situation may emerge when the derived (subjective) decision has a strong effect on the initial objective risk and can potentially induce a self-containing process of a virtual risk. The true reason of this virtual risk is the validity of data that drives the subjective perception of the perceived safety and safety loop.

In this paper, we describe the quantification of the perceived safety and perceived anxiety. The approach uses traditional 2 × 2 tables and combines this information with the Protection Motivation Theory (2), the concept of High Reliability Organizations (3), and the model of the Safety loop (5, 6) to propose a strategy on how to use scientific data for political decisions.

## Methods

### The examples used for application of the theory

We used data from different scenarios to confirm our algorithm on the quantitative assessment of ‘Perceived Safety’ and Perceived Anxiety’ using ten examples. Example #1 shows data from mammography screening reported by the Breast Cancer Surveillance Consortium (https://tools.bcsc-scc.org/BC5yearRisk/calculator.htm). The analysis considered expected outcomes for 5000 women age of forty years who are screened for breast cancer. Based on data from Carney and colleagues (8) it assumes the prevalence of breast cancer is 2 per 1000 with a mammography sensitivity of 0.66 and a specificity of 0.91. Examples #2 - #4 show data on prostate cancer screening reported by Hugosson et al. (9) when using three different endpoints. Example #2 confirms the diagnosis of prostate cancer, example #3 the disease specific mortality, and example #4 the all-cause mortality. Example #5 depicts data of a Bavarian study on mortality after myocardial infarction (10). The examples #6 - #9 are based on data reported by the Robert Koch Institute, Berlin on the Covid-19 Pandemic (11). As these reports did not include sensitivity and specificity, we used four possible combinations of sensitivity and specificity for our calculations. Example #10 uses data from the Covid-19 Pandemic Norway database FHI (Norwegian Institute of Public Health) and a report in Norwegian TV (12).

### The application of the theory

Our theory assumes that both the objective risks and the subjective perception of objective risks can be calculated from traditional 2 × 2 tables. The traditional 2 × 2 table starts with two hypotheses, e.g. a positive mammography confirms breast cancer while a negative mammography excludes breast cancer. For confirmation of the true diagnosis a gold standard, e.g. the histopathologic examination of the suspected lesion is necessary. The specimen can be collected by a fine needle biopsy. The 2 × 2 table enables the calculation of the positive and negative Likelihood Ratios (+LR; -LR). A LR value of 1 means the probability of confirmed (excluded) disease is identical in persons with a positive and with a negative test result. In other words, a test result LR = 1 is inconclusive. LR > 1, named +LR, can indicate the confirmation of a condition while LR < 1, named –LR, can indicate the exclusion of a condition. Further details for calculation and interpretation of LRs are described in Supplement I.

The calculation of the subjective perception of the objective risks follows exactly the same rules like the calculation of the objective risks in a 2 × 2 table but with exchanged X- and Y-axes. The 2 × 2 table with inverted axes starts with the communication of the test result and presumes that the communication of bad news (the bad news may be right or false) can induce ‘perceived anxiety’ while good news (independently of right or false) can induce ‘perceived safety’. The induction of perceived anxiety can be quantified by calculation of the LRs from an inverted table of a test that investigates bad news such as a diagnosis of cancer. A calculated +LR > 1 (-LR < 1) of a test that investigates the effects of bad news confirms (excludes) perceived anxiety. Correspondingly, a calculated +LR > 1 (-LR < 1) of a test that investigates the effect of good news such as prolongation of survival confirms (excludes) perceived safety.

## Results

The example shown in Table 1 uses data from the US Breast Cancer Surveillance Consortium Task Force. The summaries of all Likelihood Ratios describing the confirmation or exclusion of the investigated endpoint and of the Perceived Anxiety and Perceived Safety are shown in Table 2.

**Table 1:**
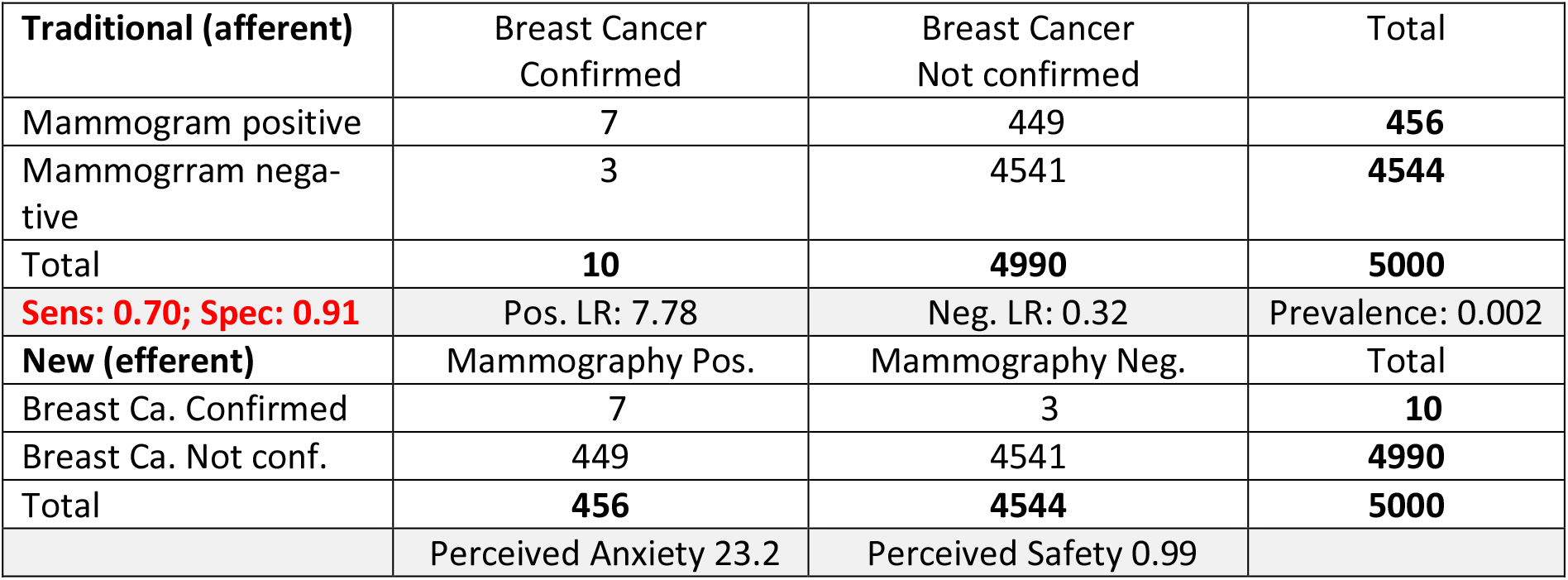
Data on example #1 breast cancer screening reported by the Breast Cancer Surveillance Consortium tool (see https://tools.bcsc-scc.org/BC5yearRisk/calculator.htm) to estimate confirmation (or exclusion) of the suspected diagnosis by calculation of the positive (or negative) Likelihood Ratio derived from the traditional 2 × 2 table. The new version (with exchanged X- and Y-axes) of the same table are used for quantification of the Perceived Anxiety (or Perceived Safety) by estimating the positive (or negative) Likelihood Ratio.

| **Traditional (afferent)** | Breast Cancer Confirmed | Breast Cancer Not confirmed | Total |
| --- | --- | --- | --- |
| Mammogram positive | 7 | 449 | **456** |
| Mammogram negative | 3 | 4541 | **4544** |
| Total | **10** | **4990** | **5000** |
| **Sens: 0.70; Spec: 0.91** | Pos. LR: 7.78 | Neg. LR: 0.32 | Prevalence: 0.002 |
| **New (efferent)** | Mammography Pos. | Mammography Neg. | Total |
| Breast Ca. Confirmed | 7 | 3 | **10** |
| Breast Ca. Not conf. | 449 | 4541 | **4990** |
| Total | **456** | **4544** | **5000** |
|  | Perceived Anxiety 23.2 | Perceived Safety 0.99 |  |

**Tab. 2:**
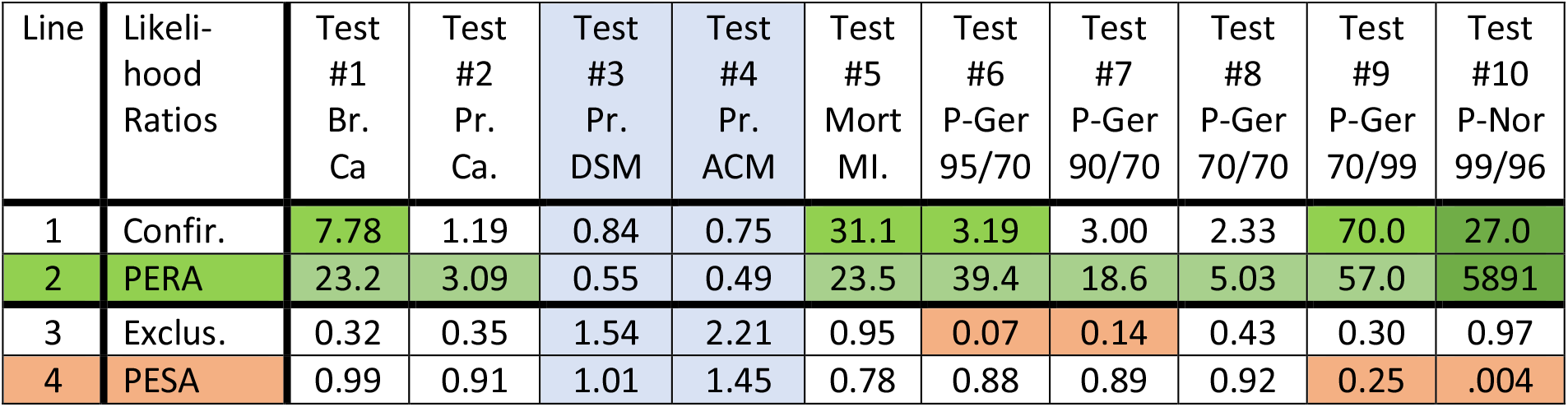
Likelihood Ratios of ten tests. Tests #3 and #4 (blue background) describe a *wanted* effect i.e. the *reduction* of Disease Specific Mortality (DSM) or of All-Cause Mortality (ACM). All other tests describe *not wanted* conditions such as evidence for Breast Cancer (Br.Ca), Prostate Cancer (PR.Ca), Mortality following myocardial infarction (mort MI), or Positive Polymerase Chain Reaction German Test or Norwegian Test including sensitivity and specificity. The positive Likelihood Ratios describe the confirmation (+LR > 3) if *not wanted conditions* are investigated in line 1 and the Perceived Anxiety in line 2 (tests #1, #2, #5, #6, #7, #8, #9, and #10). In tests #3 and #4 wanted conditions are described. The -LRs < 1 in lines 1 and 2 express the direction of the test towards exclusion of the condition and the +LR > 1 in lines 3 and 4 express the direction of the test towards confirmation of the condition. The results of tests #3 and #4 can neither confirm nor exclude an investigated condition nor perception as none of the calculated LR did exceed the limits of the indifference zone.

| Line | Likelihood Ratios | Test #1 Br. Ca | Test #2 Pr. Ca. | Test #3 Pr. DSM | Test #4 Pr. ACM | Test #5 Mort MI. | Test #6 P-Ger 95/70 | Test #7 P-Ger 90/70 | Test #8 P-Ger 70/70 | Test #9 P-Ger 70/99 | Test #10 P-Nor 99/96 |
| --- | --- | --- | --- | --- | --- | --- | --- | --- | --- | --- | --- |
| 1 | Confir. | 7.78 | 1.19 | 0.84 | 0.75 | 31.1 | 3.19 | 3.00 | 2.33 | 70.0 | 27.0 |
| 2 | PERA | 23.2 | 3.09 | 0.55 | 0.49 | 23.5 | 39.4 | 18.6 | 5.03 | 57.0 | 5891 |
| 3 | Exclus. | 0.32 | 0.35 | 1.54 | 2.21 | 0.95 | 0.07 | 0.14 | 0.43 | 0.30 | 0.97 |
| 4 | PESA | 0.99 | 0.91 | 1.01 | 1.45 | 0.78 | 0.88 | 0.89 | 0.92 | 0.25 | .004 |

Table 1 confirms fair reliability of the test for confirmation of the diagnosis of breast cancer (+LR = 7.78) but not for exclusion of the diagnosis (-LR = 0.32). The inverse table in the lower part of Table 1 shows that the +LR is highly reliable to confirm considerable Perceived Anxiety +LR = 23.2 while a negative LR, -LR = 0.99 cannot exclude Perceived Anxiety. The corresponding data from additional nine scenarios are shown in Supplement II.

The summary results (Table 2) of all ten scenarios described in lines 1 and 3, the LRs that express the functions of a test either to confirm (line 1) or to exclude (line 3) the investigated conditions. Lines 2 and 4 describe the LRs that either confirm (line 2) or exclude (line 4) the induction of perceived anxiety (PERA; line 2) or perceived safety (PESA; line 4).

In line 1 eight of the ten tests show a +LR > 1 but two tests (tests #3 and #4) show a -LR < 1. The positive LRs confirm the investigated conditions while the negative LRs exclude the investigated condition. To understand the results of 2 × 2 tables it is necessary to consider the valence of confirmation (good news or bad news) for correct interpretation of the results. The eight tests that generated +LR assumed bad news, e.g. death or infection whereas the two tests (test #3 and #4) that generated -LR, expressed good news i.e. reduction of mortality.

Line 1 of Tab. 2 shows that three tests (test #5, #9, #10) confirm very likely (+LR > 10) the investigated conditions, two additional tests (test #1 and #6) confirm likely (+LR > 3) the investigated condition, and five tests (tests #2, #3, #4, #7, and #8) can neither confirm nor reject the investigated condition because their LRs do not exceed the likelihood indifference zone (LRs between 0,3 and 3). The three tests (tests #2, #7, #8) with +LR > 1 failed to confirm the investigated condition and two tests (tests #3 and #4) with -LR > 0.3 failed to exclude the investigated condition.

Line 2 of Table 2 describes that eight of the ten tests result in increased anxiety. One of these tests (test #10) shows a very high probability on increased anxiety by a +LR > 3000. Five tests (test #1, #5, #6, #7, #9) confirm very likely the induction of anxiety by a +LR > 10. Two additional tests (tests #2 and #8) confirm moderately the induction of anxiety by a +LR > 3, but two other tests (test #3 and #4) fail to exclude the induction of anxiety by -LRs of 0.55 and 0.49.

Line 3 of Table 2 shows that a -LR < 0.3 that would be strong enough to exclude the investigated condition, was observed in only two of ten tests (test #6 and #7). These two tests exclude the presumed correlation of PCR with viral disease.

Line 4 shows that two of the ten test exclude the induction of perceived safety (PERA) (i.e. test #9 and #10). The remaining eight tests can neither confirm nor exclude the perception of safety.

The LRs in Table 2 also demonstrate that the traditional tables (description of cases detected) and the inverted tables (description of the induced psychological perception) generate different and independent results.

Figure 2 provides a graphical logarithmic presentation of the data. It shows the correlation between the objective functions (X-axis expressed as exclusion or confirmation) of tests and the subjective perception of the objective functions (Y-axis expressed as Perceived Safety or Perceived Anxiety). Most tests cannot exclude (blue points on X-axis) but can confirm (yellow points on X-axis) a diagnosis. Accordingly, most of our investigated tests cause Perceived Anxiety (PERA) but not Perceived Safety (PESA).

**Fig 2:**
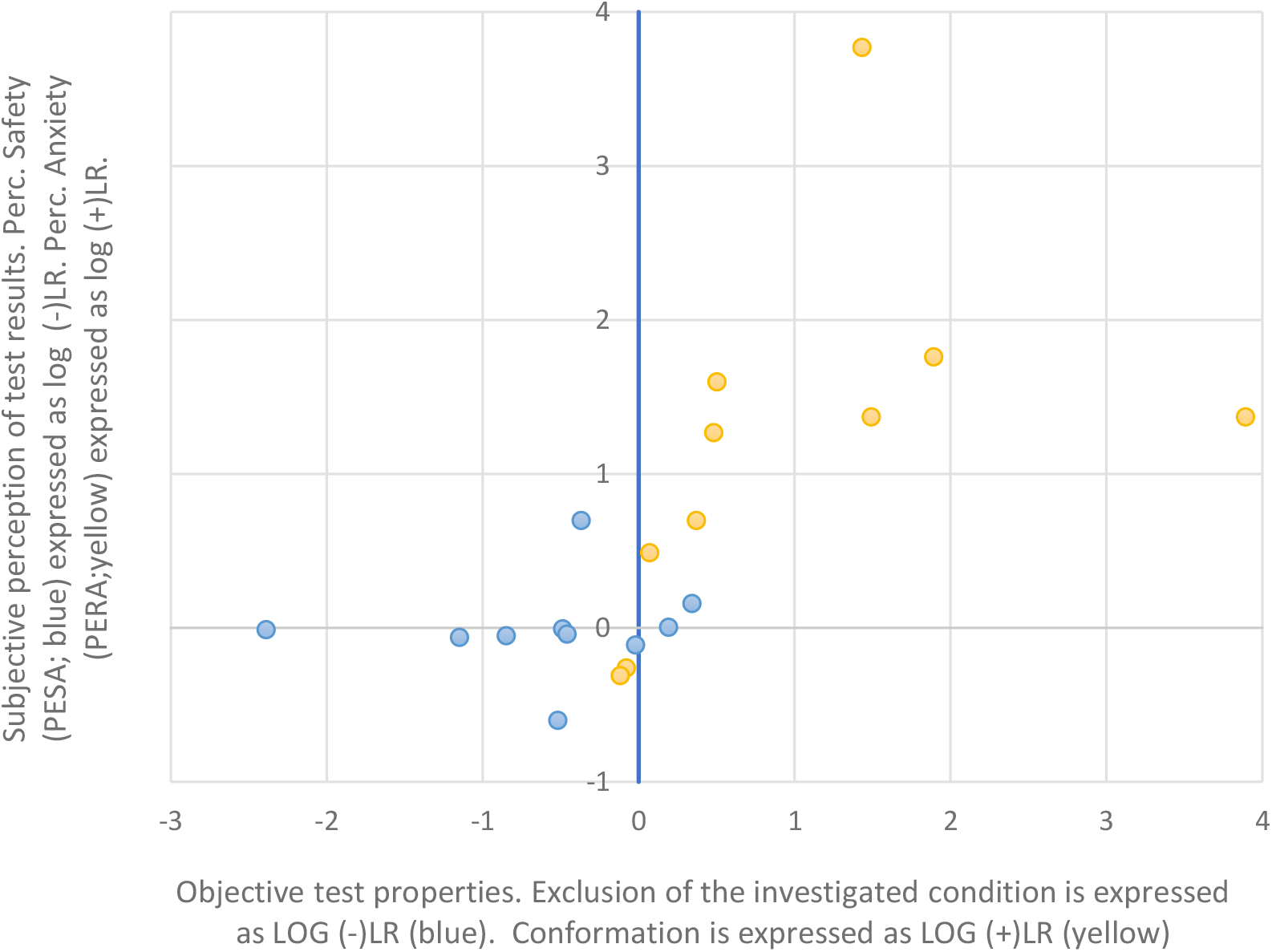
Correlation of the objective functions (X-axis expressed as exclusion or confirmation) of tests and the subjective perception of the objective functions (Y-axis expressed as Perceived Safety or Perceived Anxiety) caused by these tests.

## Discussion

Our results demonstrate that each 2 × 2 test table has two independent sets of functions. The first set of (traditional) functions describes the correlation of a test result with a confirmed diagnosis. The second set of (new) functions describes the correlation of the confirmed diagnosis with the generation of psychological effects. Our results also show that any inappropriate interpretation of a test result will induce harmful effects that may cause - via an incorrect diagnosis – an inappropriate treatment of a single patient or may cause - via inappropriate political decision - a national or even international policy failure.

Due to the strong psychological effects that can be caused by inappropriate interpretation of test results, it is important to distinguish between professional consideration and public communication of test results and their interpretations. Some examples may be useful to elucidate the ‘explosive force’ of test results.

Tests describe only probabilities, never certainty. Gerd Gigerenzer and colleagues have published material for experts and ‘ordinary people’ to guide interpretation of numbers, tests, probabilities, and conditional probabilities (13, 14). Understanding the statistical principles underlying tests is the first of two necessary steps. The second step is the derivation of correct decisions based on test results. The correct derivation of practical consequences requires different knowledge than the correct interpretation of statistical test result.

Mammography screening or a polymerase chain reaction (PCR) or an antibody test provide instructive examples. A positive mammography test does not mean the woman suffers from breast cancer and a positive PCR or antibody test does not confirm a viral disease. Tests are imperfect and any positive test result may be correct or false. Elimination of the false positive test results requires a gold standard of an indicator of “ground truth”. In case of mammography the gold standard is the histopathologic confirmation of the diagnosis in a specimen taken by a fine needle biopsy. The diagnosis of viral infection is more difficult as there are very mild courses of the disease that are sometimes not even recognized as disease. On the other hand, it is known that elderly and multimorbid people belong to high risk groups in a pandemic. It does not make sense to request the use of not validated tests because nobody can estimate the true risk of a person with a positive PCR test result to be sick and the chance of a person with a negative PCR test result to be healthy. To generate reliable data, we need validated tests for both the detection of viral particles and the detection of antibodies against specific viral structures (15).

We strongly advice to consideration of uncertainty in the interpretation of test results. Expert knowledge has improved, and new research will continue to modify test interpretation. By combining the concept of the Safety Loop with the ‘Protection Motivation Theory the ‘Theory of a Safety-Protection-Loop’ (Fig. 3) may provide a valuable tool for guiding communication between providers and patients.

**Fig. 3:**
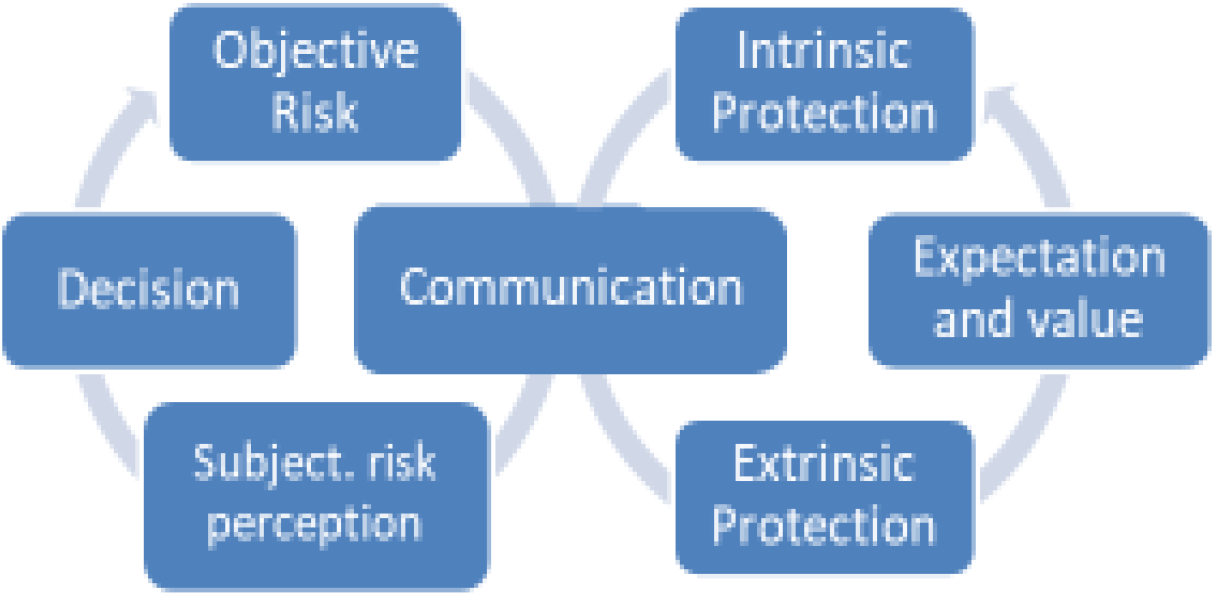
The Safety-Protection Loop combines two loops. The first loop describes the association of objective risks via communication with the subjective perception of the objective risk (‘perceived safety’). The second loop illustrates the modification of the communication by the Protection Motivation Theory that includes both the extrinsic protection (represented by exposition to external information) that influences the Intrinsic Protection (defined by the individual personality) via the modulation of expectations and value. Both loops meet at the level of communication.

The interface between medicine and public policy remains challenging. Medical professionals are usually not trained in public policy and policy makers are rarely qualified medical doctors. We may need a new professional group that can bridge medicine and politics. Controversial discussions of results are essential in science but may be disturbing for political decisions. The new profession of investigative journalists might get advanced training in the communication of conflicting scientific perspectives. They should identify the published peer reviewed knowledge, to exclude immature considerations and opinions, to moderate the controversial scientific discussion on solid data, and finally extract the currently reliable state of the art from a dynamic scientific process. This process could be open to the public and may immediately be available for the policy discussions. The goal of any consequences and the assessed endpoints and time intervals should be defined to restore the lost confidence in the policy decision process.

## Data Availability

Data has been reported in published articles or is included in the article as supplementary material

## Supplement I: Examples for calculation of the +LR & -LR and of PERA & PESA

The upper part of the example explains the calculation of +LR & -LR. The lower part of the example shows the exchanged X- and Y-axes of the same table as above. By using the version of the table with exchanged axes the calculation of PERA & PESA can be completed by the same strategy that was used for calculation of the +LR & -LR.

The result of the (+)LR describes the x-fold excess of the frequency of a positive test in the group with the conditions (e.g. in women with biopsy confirmed breast cancer) than in the group without this condition (i.e. no biopsy confirmed breast cancer).

Please remember each test has two different types of qualities. A test can either confirm an assumption of an *unwanted condition* or reject it or both ore none. The +LR confirms an unwanted condition. +LR = 5 means the chance to find a test-positive person among those with the unwanted condition will be 5-times higher than the chance to find a test-positive person among those without this condition.

LR = 1 means neither confirmation nor exclusion. The closer the LR is to the value of 1 the higher is the risk of this test neither to confirm nor to exclude. Therefore, many researchers consider results in the zone around LR = 1, i.e. between a -LR of 0.3 and a +LR of 3.0 a zone of indifference. -LR between 0.3 and 0.1 can be considered fairly good to exclude a condition and those smaller than 0.1 can almost certainly exclude a condition. Corresponding results apply to +LR. +LR between 3.0 and 10.0 may fairly well confirm a condition and values above 10.0 almost certainly confirm a condition.

We are using the example of breast cancer to demonstrate that the quality of the ‘Gold Standard’ may vary depending on the type of the selected ‘Gold Standard’. One of the excellent Gold Standards for breast cancer is the confirmation of advanced stage of disease because even biopsy confirmed breast cancer may disappear spontaneously (Kaplan et al. ; Porzsolt et al.) or will not be detected by mammography in 3 of 10 cases (see supplement II). Such residual uncertainties should be considered in any type of test. The following 2 × 2 table is a standard version for confirmation of both LRs:

| | Condition met | Condition unmet | $\Sigma$ |
| --- | --- | --- | --- |
| Test + | A | B | A + B |
| Test - | C | D | C + D |
| $\Sigma$ | A + C | B + D | A + B + C + D |

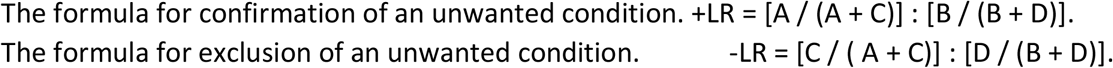

The corresponding table with exchanged X- and Y-axes for calculation of PERA & PESA is:

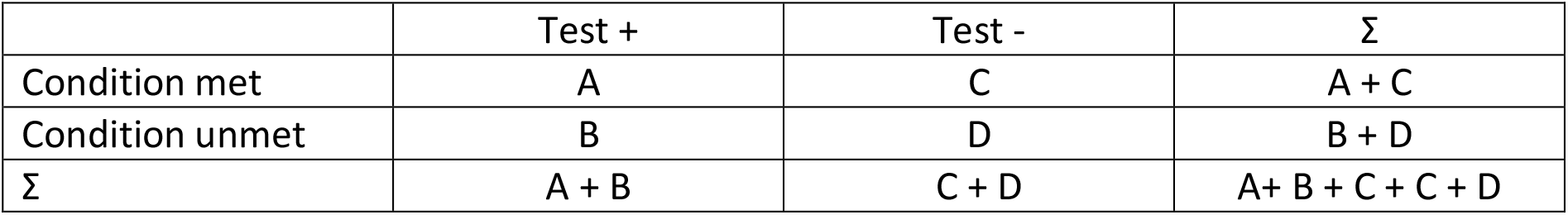

The formula for confirmation of the effect of the unwanted condition i.e. Perceived Anxiety (PERA) is

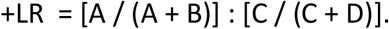

The formula for exclusion of the effect of the unwanted condition i.e. Perceived Safety (PESA)is

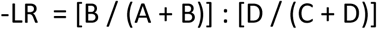

## Supplement II

Examples #2 - #10. The following tables show the raw data and calculated values summarized in Tab. 2 of the related publication.

Example #2 Prostate Cancer Screening (Endpoint Cancer) (Ref: Hugosson)

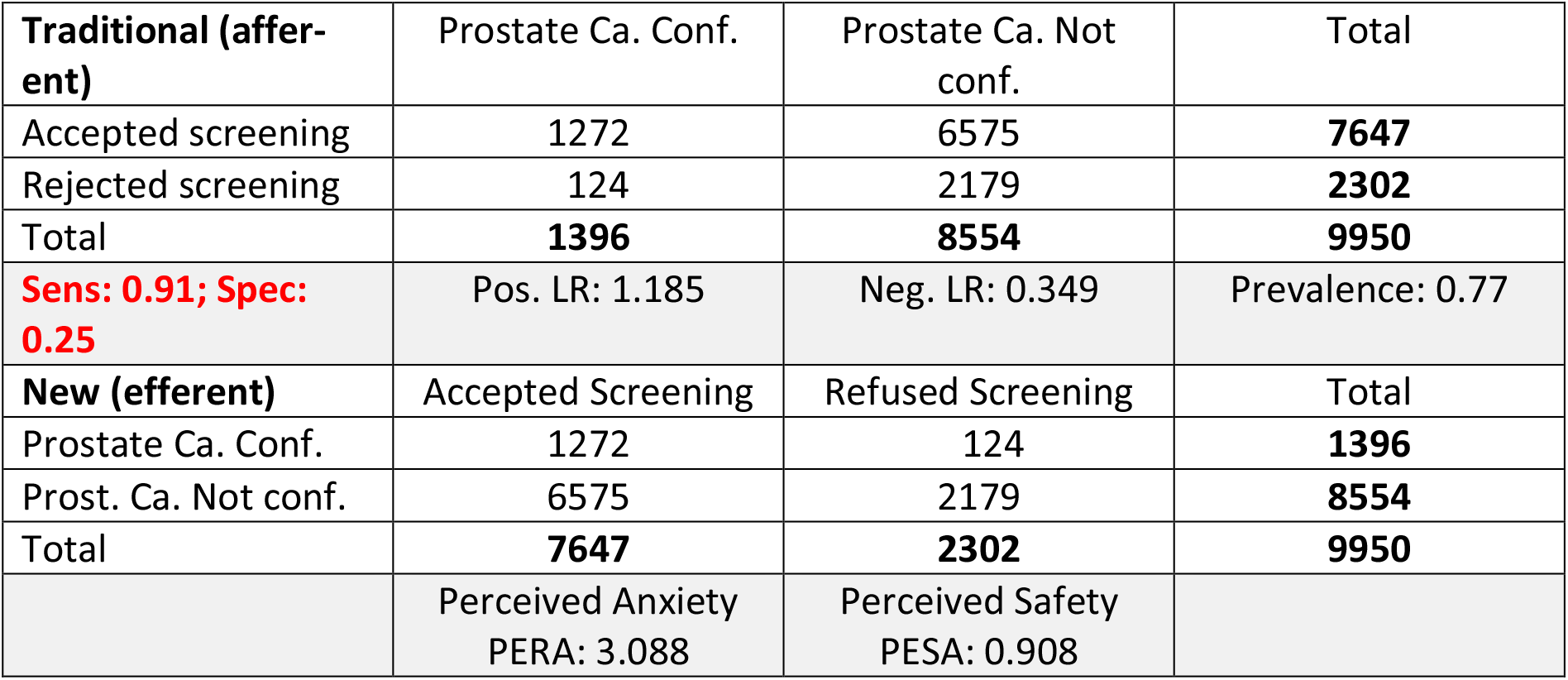

Example #3 Prostate Cancer Screening (Endpoint Disease Specific Mortality) (Ref: Hugosson)

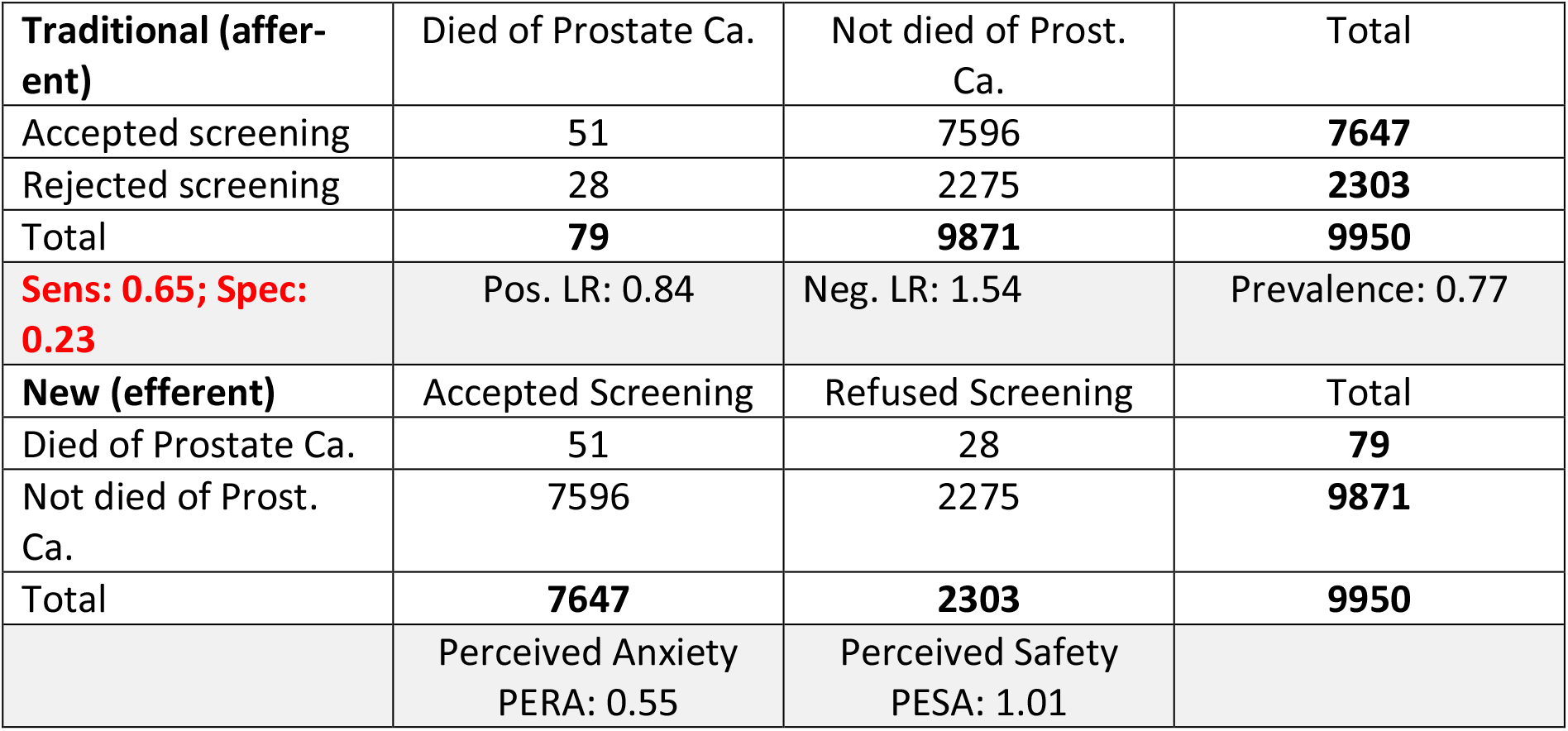

Example #4 Prostate Cancer Screening (Endpoint All-cause Mortality) (Ref: Hugosson)

| **Traditional (affer-ent)** | Dead (any cause) | Alive | Total |
| --- | --- | --- | --- |
| Accepted screening | 1763 | 5884 | **7647** |
| Rejected screening | 1081 | 1222 | **2303** |
| Total | **2844** | **7106** | **9950** |
| Sens: 0.62; Spec: 0.17 | Pos. LR: 0.75 | Neg. LR: 2.21 | Prevalence: 0.77 |
| **New (efferent)** | Accepted Screening | Refused Screening | Total |
| Dead (any cause) | 1763 | 1081 | **2844** |
| Alive | 5884 | 1222 | **7106** |
| Total | **7647** | **2303** | **9950** |
|  | Perceived Anxiety<br>PERA: 0.49 | Perceived Safety<br>PESA: 1.45 |  |

Example #5 Survival following Myocardial Infarction (MI) x10E5 (Ref: Bavarian Ministry Social Affairs)

|  |  |  |  |
| --- | --- | --- | --- |
| **Traditional (affer-ent)** | Heart related death | Death other reason | Total |
| With MI | 0.069 | 0.231 | **0.3** |
| Without MI | 1.27 | 128.43 | **129.7** |
| Total | **1.34** | **128.66** | **130.0** |
| **Sens: 0.05; Spec: 1.00** | Pos. LR: 28.68 | Neg. LR: 0.95 | Prevalence: 0.002 |
| **New (efferent)** | With MI | Without MI | Total |
| Heart related death | 0.069 | 1.27 | **1.34** |
| Death other reason | 0.231 | 128.43 | **128.66** |
| Total | **0.300** | **129.70** | **130.00** |
|  | Perceived Anxiety<br>PERA: 23.5 | Perceived Safety<br>PESA: 0.78 |  |

Example #6 Pandemic PCR-Test. (Ref: Robert Koch Institute Berlin / Germany)

|  |  |  |  |
| --- | --- | --- | --- |
| **Traditional (affer-ent)** | Disease Spec. Mor-tal. | Alive | Total |
| Pat. with positive test | 6646 | 46993 | **53639** |
| Without positive test | 350 | 110818 | **111168** |
| Total | **6996** | **157811** | **164807** |
| **Sens: 0.95; Spec: 0.70** | Pos. LR: 3.19 | Neg. LR: 0.07 | Prevalence: 0.33 |
| **New (efferent)** | Pat with positive test | Pat without pos. test | Total |
| Disease Spec. Mor-tal. | 6646 | 350 | **6996** |
| Alive | 46993 | 110818 | **157811** |
| Total | **53639** | **111168** | **164807** |
|  | Perceived Anxiety<br>PERA: 39.4 | Perceived Safety<br>PESA: 0.88 |  |

Example #7 Pandemic PCR-Test. (Ref: Robert Koch Institute Berlin / Germany)

| **Traditional (affer-ent)** | Disease Spec. Mor-tal. | Alive | Total |
| --- | --- | --- | --- |
| Pat. with positive test | 6296 | 47343 | **53639** |
| Without positive test | 700 | 110468 | **111168** |
| Total | **6996** | **157811** | **164807** |
| Sens: **0.90**; Spec: **0.70** | Pos. LR: 3.00 | Neg. LR: 0.14 | Prevalence: 0.33 |
| **New (efferent)** | Pat with positive test | Pat without pos. test | Total |
| Disease Spec. Mor-tal. | 6296 | 700 | **6996** |
| Alive | 47343 | 40468 | **157811** |
| Total | **53639** | **111168** | **164807** |
|  | Perceived Anxiety<br>PEAN: 18.6 | Perceived Safety<br>PESA: 0.89 |  |

Example #8 Pandemic PCR-Test. (Ref: Robert Koch Institute Berlin / Germany)

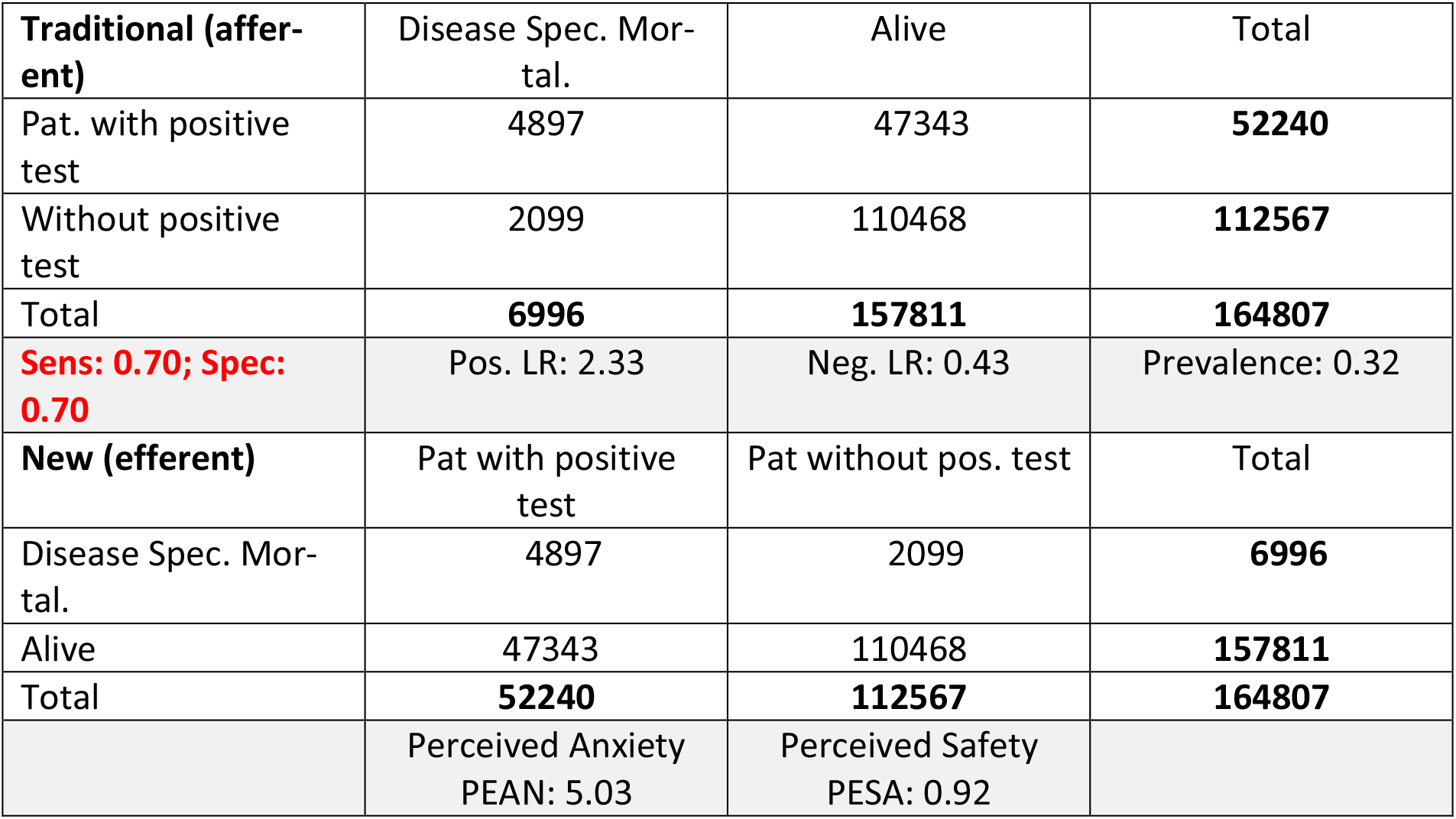

| **Traditional (affer-ent)** | Disease Spec. Mor-tal. | Alive | Total |
| --- | --- | --- | --- |
| Pat. with positive test | 4897 | 47343 | **52240** |
| Without positive test | 2099 | 110468 | **112567** |
| Total | **6996** | **157811** | **164807** |
| Sens: **0.70**; Spec: **0.70** | Pos. LR: 2.33 | Neg. LR: 0.43 | Prevalence: 0.32 |
| **New (efferent)** | Pat with positive test | Pat without pos. test | Total |
| Disease Spec. Mor-tal. | 4897 | 2099 | **6996** |
| Alive | 47343 | 110468 | **157811** |
| Total | **52240** | **112567** | **164807** |
|  | Perceived Anxiety<br>PEAN: 5.03 | Perceived Safety<br>PESA: 0.92 |  |

Example #9 Pandemic PCR-Test. (Ref: Robert Koch Institute Berlin / Germany)

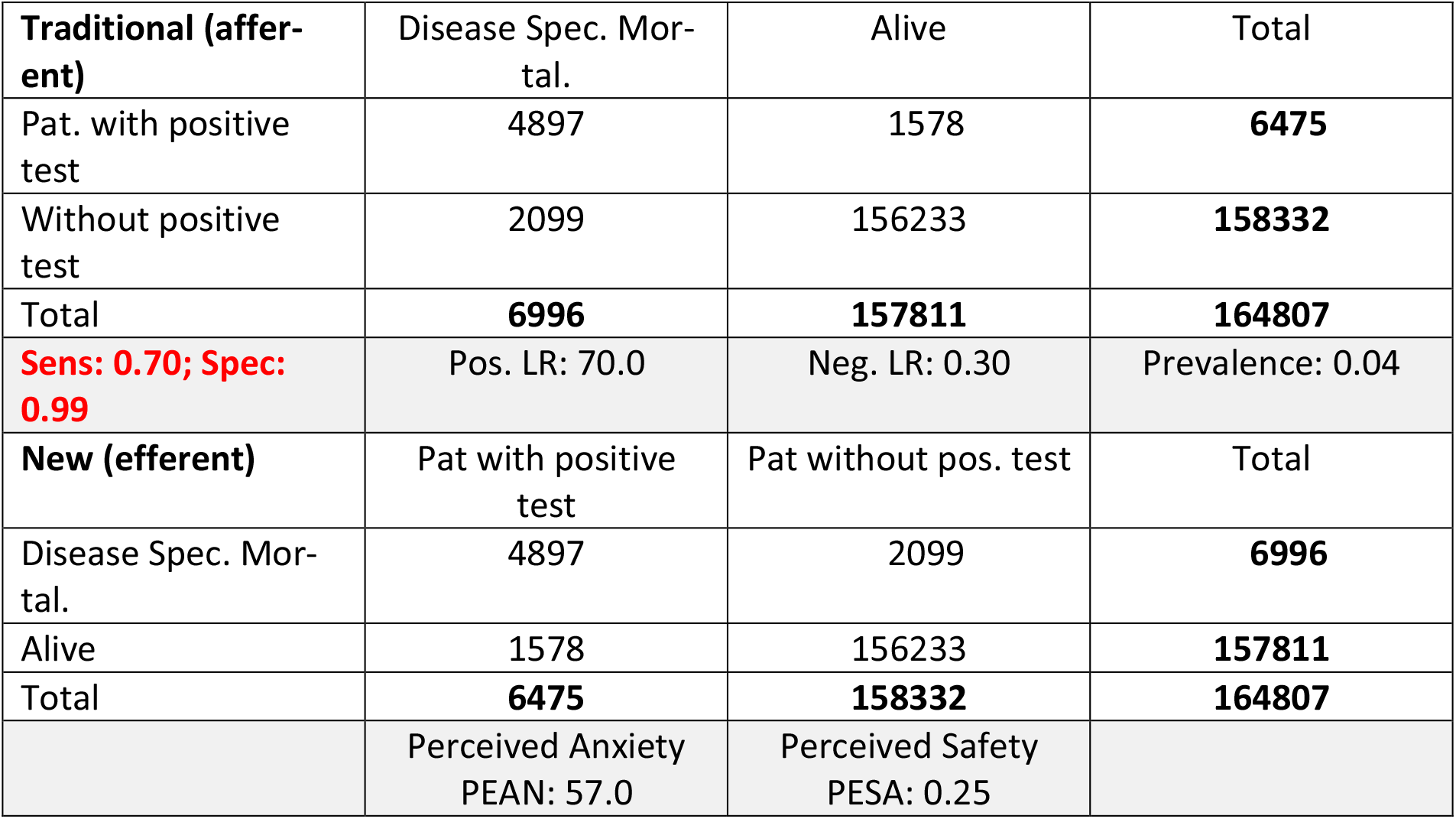

| **Traditional (affer-ent)** | Disease Spec. Mor-tal. | Alive | Total |
| --- | --- | --- | --- |
| Pat. with positive test | 4897 | 1578 | **6475** |
| Without positive test | 2099 | 156233 | **158332** |
| Total | **6996** | **157811** | **164807** |
| **Sens: 0.70; Spec: 0.99** | Pos. LR: 70.0 | Neg. LR: 0.30 | Prevalence: 0.04 |
| **New (efferent)** | Pat with positive test | Pat without pos. test | Total |
| Disease Spec. Mor-tal. | 4897 | 2099 | **6996** |
| Alive | 1578 | 156233 | **157811** |
| Total | **6475** | **158332** | **164807** |
|  | Perceived Anxiety<br>PEAN: 57.0 | Perceived Safety<br>PESA: 0.25 |  |

Example # 10 Pandemic PCR-Test. (Ref: **FHI (May 16,2020)** & NRK via Arctic University of Tromsø/Norway)

| **Traditional (affer-ent)** | Disease Spec. Mor-tal. | Recovered | Total |
| --- | --- | --- | --- |
| Pat. with positive test | **232** | 7965 | **8197** |
| Without positive test | 1 | 208152 | 208153 |
| Total | 233 | 216117 | **216350** |
| **Sen: 0.996 Spec: 0.963** | Pos. LR: 27.0 | Neg. LR: 0.004 | Prevalence: 0.038 |
| **New (efferent)** | Pat with positive test | Pat without pos. test | Total |
| Disease Spec. Mor-tal. | 232 | 1 | 233 |
| Recovered | 7965 | 208152 | 216117 |
| Total | **8197** | 208153 | **216350** |
|  | Perceived Anxiety<br>PERA: 5891.3 | Perceived Safety<br>PESA: 0.97 |  |

